# Impact of Dialysis Duration and Frailty on Frailty Progression: A Japanese Nationwide Cohort Study

**DOI:** 10.1101/2025.06.04.25328972

**Authors:** Kakuya Niihata, Noriaki Kurita, Ryohei Inanaga, Tatsunori Toida, Masanori Abe, Takao Masaki, Suguru Yamamoto

## Abstract

**Importance:** Frailty is common among dialysis patients and significantly affects the quality of life for both patients and their caregivers. However, limited evidence exists on the long- term changes in physical function in this population.

**Objective:** To examine 8-year trajectories of physical function and assess associations with baseline dialysis duration and physical function status among Japanese dialysis patients.

**Design:** Nationwide, cohort study.

**Setting:** Japan; data obtained from a registry.

**Participants:** 223,501 Japanese adults receiving hemodialysis enrolled in the 2010 Japanese Society for Dialysis Therapy Renal Data Registry.

**Exposures:** Baseline dialysis duration (<5, 5–<10, 10–<20, 20–<30, ≥30 years) and physical function were assessed using the Eastern Cooperative Oncology Group Performance Status, categorized as non-frail, frail, or bedridden.

**Main Outcomes and Measures:** Physical function at 8 years was similarly classified as non- frail, frail, bedridden, or deceased. Multinomial logistic regression was used to estimate adjusted odds ratios, average marginal effects, and predicted probabilities based on baseline exposures.

**Results:** Among patients with complete baseline and 8-year follow-up data, 59.9% died, 8.8% became frail, 2.4% were bedridden, and 28.9% remained non-frail. Longer dialysis duration and baseline frailty or bedridden status were associated with increased odds of subsequent frailty, bedridden status, and mortality. Compared with patients with <5 years of dialysis, those with ≥30 years had a 1.6% (95% confidence interval, 0.6%–2.6%) higher probability of frailty and a 13.0% (95% confidence interval, 11.8%–14.3%) higher probability of death. Compared with non-frail status at baseline, frailty was associated with a 0.0% (95% confidence interval, −0.4% to 0.4%) change in frailty and a 15.8% (95% confidence interval, 14.5%–17.0%) increase in death; bedridden status was associated with a 1.7% (95% confidence interval, 1.1%–2.3%) increase in being bedridden and a 27.6% (95% confidence interval, 26.5%–28.8%) increase in death.

**Conclusions and Relevance:** In this nationwide 8-year study, majority of the hemodialysis patients experienced either death or functional decline. Longer dialysis duration and baseline frailty were associated with adverse outcomes, although absolute increases in frailty were modest. These findings highlight the need for early, values-based shared decision-making in the management of dialysis patients.

## Introduction

Frailty, characterized by increased vulnerability, is common among dialysis patients, with a meta-analysis reporting a rate of 34.3%^1^ and our nationwide study of 230,000 Japanese patients identifying a prevalence of 36.2%.^2^ Among patients undergoing dialysis, particularly older adults, independence is often prioritized over survival.^3^ Patients and caregivers highly value outcomes such as reduced fatigue and the ability to travel or work.^4^ Therefore, preventing the onset or progression of frailty is a critical concern. In addition, frailty affects the daily quality of life of both patients and their caregivers. Evidence suggests that declines in physical function and increases in symptom burden, both closely linked to frailty, can increase the burden on informal caregivers and lead to feelings of guilt among patients.^5,6^ To promote patient-centered care that aligns with the priorities of patients and families, patients must be informed about how frailty evolves during dialysis therapy.^7,8^ Despite advances in dialysis care that enable for long-term treatment,^2,8^ evidence remains limited on how frailty transitions evolve over time with increasing dialysis duration and differing baseline frailty levels.

A previous study investigating frailty trajectories in hemodialysis (HD) patients has been limited by short follow-up durations (up to two years) and a lack of dialysis duration data.^9^ Another study examined frailty changes over nearly four years in peritoneal dialysis (PD) patients; however, a small sample size (<300) could not demonstrate the impact of baseline frailty.^10^ Our prior study, based on data from the Japanese Society for Dialysis Therapy Renal Data Registry (JRDR), comprising 230,000 Japanese patients, found a higher prevalence of frailty among those with longer dialysis durations, particularly those undergoing treatment for >30 years.^2^ Nevertheless, because the study was cross-sectional, it could not elucidate longitudinal changes in frailty or assess the influence of dialysis duration on those changes.

Notably, frailty index data collected by the JRDR eight years before the 2018 dataset we analyzed provides a unique opportunity to examine long-term frailty transitions.

Therefore, we conducted a nationwide cohort study of Japanese dialysis patients using the JRDR database to describe 8-year trajectories of frailty and other physical function statuses, and to evaluate the impact of baseline dialysis duration and physical function on outcomes after 8 years. The findings are expected to provide essential information for improving life planning focused on patient- and caregiver-centered priorities, and to support individualized care planning during dialysis and informed decision-making prior to dialysis initiation.^7,8^

## Methods

### Study design and participants

This cohort study utilized anonymized data from the JRDR database, which compiles annual nationwide data from nearly all dialysis facilities in Japan. Detailed descriptions of the JRDR have been previously reported.^11^ For the current study, we defined baseline data as those collected at the end of 2010. According to the Japanese Society for Dialysis Therapy (JSDT), the 2010 survey achieved a response rate of 98.6% (4,166/4,226 facilities).^12^ Physical function status was tracked using follow-up data from patients registered at the end of 2018, and mortality outcomes were monitored annually from 2010 to 2018.

Patients were included if they (1) were aged 20 years or older and (2) received HD, hemodiafiltration, or hemofiltration. Exclusion criteria comprised the following: (1) received only PD or had an unknown treatment status in 2010, (2) had missing outcome data, and (3) underwent kidney transplantation or discontinued dialysis between 2010 and 2018. Based on previous studies,^13^ outliers were treated as missing values if they met any of the following criteria: (1) serum phosphorus <0.5 mg/dL, (2) height <80 cm or >200 cm, (3) body weight <20 kg or >150 kg, (4) serum creatinine <3.0 mg/dL or >20 mg/dL, (5) blood urea nitrogen <10 mg/dL or >250 mg/dL, (6) single-pool Kt/V <0.5 or >4.0, (7) β2-microglobulin (β2MG) <5.0 mg/dL or >100 mg/dL, (8) serum albumin <0.5 mg/dL or >5.0 mg/dL, (9) serum C-reactive protein (CRP) >50 mg/dL, (10) hemoglobin <5.0 g/dL or >20 g/dL, and (11) normalized protein catabolic rate <0.3 g/kg/day or >2.0 g/kg/day.

### Ethical considerations

The JRDR survey was conducted in accordance with the Japanese Ethical Guidelines for Epidemiological Studies published by the Ministry of Education, Science and Culture, and the Ministry of Health, Labor, and Welfare.^14^ The study protocol was approved by the Ethics Committee of the JSDT (Approval No.68), and the study was conducted in accordance with the tenets of Declaration of Helsinki. The need for informed consent was waived because the dataset contained no identifiable information.

### Outcome

The primary outcome was a four-category classification of physical function status at the 8- year follow-up (2018): non-frail, frail (excluding bedridden status), bedridden, and dead.

Non-frail status corresponded to an Eastern Cooperative Oncology Group Performance Status (ECOG PS) grade of 1.^15,16^ Frailty included grades 2 and 3, while bedridden status corresponded to grade 4. Grade 2, characterized by preserved self-care but impaired ability to work, has been used as a marker of frailty in previous studies and served as the threshold for this classification.^2,17^ Based on a previous study, death, considered a higher-order outcome than non-frail or frail status,^9^ was defined as any mortality recorded in annual follow-up data from 2010 to 2018.

### Primary Exposure

The primary exposures were dialysis vintage and physical function status at baseline. Dialysis vintage was categorized as follows: 0–<5, 5–<10, 10–<20, 20–<30, and ≥30 years.^2^ Physical function was assessed using the ECOG PS scale and categorized as non-frail, frail, or bedridden.

### Covariates

Covariates were defined as baseline variables potentially influencing physical function status. Selection of these covariates was guided by evidence from the literature and the clinical expertise of the investigators (KN, TT, RI, SY, NK).

The following variables were selected as potential covariates: age, sex, body mass index, serum albumin, corrected serum calcium, serum phosphorus, intact parathyroid hormone (PTH), hemoglobin, serum C-reactive protein, β2MG, creatinine index, normalized protein catabolic rate, single-pool Kt/V, diabetes, dementia, ischemic heart disease, cerebral hemorrhage, cerebral infarction, limb amputation, carpal tunnel syndrome, history of hip fractures, and residential setting.

Corrected serum calcium was calculated as follows:

Corrected serum calcium = serum calcium + (4 − serum albumin) (if serum albumin is <4 mg/dL).

To estimate muscle mass in HD patients, the creatinine index was calculated based on a previously reported formula:^18^

Creatinine index = 16.21 + 1.12 × sex (0 for female, 1 for male) − 0.06 × age (in years) − 0.08 × single-pool Kt/V + 0.009 × 88.4 × serum creatinine (mg/dL).

Serum levels of corrected calcium, phosphorus, intact PTH, and hemoglobin were categorized into three groups based on the JSDT guidelines:^19,20^ <8.4 mg/dL, 8.4-10.0 mg/dL, >10.0 mg/dL for corrected calcium; <3.5 mg/dL, 3.5-6.0 mg/dL, >6.0 mg/dL for phosphorus; <60 pg/mL, 60-240 pg/mL, >240 pg/mL for intact PTH; and <10 g/dL, 10-12 g/dL, >12 g/dL for hemoglobin.

### Statistical analysis

Descriptive statistics are presented as medians with interquartile ranges for continuous variables and frequencies with percentages for categorical variables. To visualize the trajectory of physical function status, a Sankey diagram was used to illustrate transitions from baseline to the 8-year follow-up.^21^

The association between physical function status at 8 years (categorized as non-frail, frail, bedridden, or dead) and baseline dialysis duration and physical function status (categorized as non-frail, frail, or bedridden) was examined using a multinomial logistic regression model.

All covariates described earlier were included in the model. Adjusted odds ratios (ORs) were estimated with non-frail status as the reference outcome category (i.e., base outcome). For instance, the OR for frail status at the 8-year follow-up among individuals with ≥30 years of dialysis, compared with those with <5 years (reference exposure category), quantified the relative likelihood of being frail rather than non-frail. ^22^

Average marginal effects (AMEs) were estimated using the same model to assess absolute differences in the probability of each physical function status at 8 years based on baseline dialysis duration and physical function status.^23^ This approach enabled quantification, for example, the percentage difference in the probability of being frail at 8 years for individuals with ≥30 years of dialysis compared with those with <5 years. Predicted probabilities were also estimated using the same model and the adjusted average prediction approach to quantify the expected probability of each physical function status at 8 years. These estimates (AMEs and predicted probabilities) represent expected values in a counterfactual population, assuming variation in only the variable of interest (e.g., dialysis duration) while holding all other covariates constant.^24^

Missing data were imputed using multiple imputation by chained equations, generating 20 imputed datasets, which were combined following Rubin’s rule.^25^ Statistical significance was set at P < 0.05. All analyses were conducted using STATA (version 18; Stata LP, College Station, TX, USA).

## Results

### Patient characteristics

A total of 223,501 patients were included in the analysis. Patient selection flow and characteristics are shown in Supplementary Figure 1 and Table 1, respectively. The median age was 68 years (IQR, 60–76), and the median dialysis duration was 5 years (IQR, 2–10). The distribution of patients by dialysis vintage was as follows: 47.3% had received dialysis for 0–<5 years, 25.7% for 5–<10 years, 19.3% for 10–<20 years, 5.8% for 20–<30 years, and 1.8% for ≥30 years. At baseline, 73.0% of patients were classified as non-frail, 20.7% as frail, and 6.3% as bedridden. Patient characteristics by dialysis vintage and baseline functional status are presented in Supplementary Tables 1 and 2, respectively.

**Table 1.** Baseline characteristics (N = 223,501)

|  | **Total**<br>**N = 223,501** | **Missing, n** |
| --- | --- | --- |
| **Demographic** |  |  |
| Age | 68 [60 - 76] | 0 |
| Male sex | 78,524 (38.2%) | 0 |
| Dialysis duration |  | 89 |
| 0-<5 years | 105,729 (47.3%) |  |
| 5-<10 years | 57,488 (25.7%) |  |
| 10-<20 years | 43,180 (19.3%) |  |
| 20-<30 years | 13,044 (5.8%) |  |
| ≥30 years | 3,971 (1.8%) |  |
| BMI, kg/m<sup>2</sup> |  | 41,383 |
| 18.5-<25 | 117,276 (64.4%) |  |
| <18.5 | 40,629 (22.3%) |  |
| 25-<30 | 20,234 (11.1%) |  |
| ≥30 | 3,979 (2.2%) |  |
| Baseline functional status |  | 34,505 |
| Non-frail | 137,947 (73.0%) |  |
| Frail | 39,191 (20.7%) |  |
| Bedridden status | 11,858 (6.3%) |  |
| Residential setting |  | 31,658 |
| Home | 170,287 (88.8%) |  |
| Nursing home | 4,027 (2.1%) |  |
| Hospital | 17,529 (9.1%) |  |
| **Laboratory investigation** |  |  |
| Hb, g/dL |  | 21,093 |
| 10-<12 | 119,773 (59.2%) |  |
| <10 | 61,034 (30.2%) |  |
| ≥12 | 21,601 (10.7%) |  |
| Ca, mg/dL |  | 24,400 |
| 8.4-10.0 | 147,152 (73.9%) |  |
| <8.4 | 21,635 (10.9%) |  |
| >10.0 | 30,314 (15.2%) |  |
| P, mg/dL |  | 19,096 |
| 3.5-6.0 | 135,178 (66.1%) |  |
| <3.5 | 19,601 (9.6%) |  |
| >6.0 | 49,626 (24.3%) |  |
| intact PTH, pg/mL |  | 41,554 |
| 60-240 | 105,138 (57.8%) |  |
| <60 | 43,609 (24.0%) |  |
| >240 | 33,200 (18.3%) |  |
| Creatinine index | 20.7 [18.8 - 22.8] | 33,033 |
| Albumin, mg/dL | 3.7 [3.4 - 3.9] | 23,924 |
| CRP, mg/dL | 0.1 [0 - 0.4] | 59,229 |
| β2MG, mg/L | 26.7 [22.5 - 30.8] | 53,749 |
| Single-pool Kt/V | 1.39 [1.2 - 1.59] | 31,669 |
| nPCR | 0.86 [0.74 - 0.98] | 30,791 |
| **Cause of ESKD** |  | 17,703 |
| Diabetic kidney disease | 78,524 (38.2%) |  |
| Glomerulonephritis | 72,543 (35.3%) |  |
| Hypertensive nephrosclerosis | 17,910 (8.7%) |  |
| Polycystic kidney disease | 6,948 (3.4%) |  |
| RPGN | 1,100 (0.5%) |  |
| Others | 28,773 (14.0%) |  |
| **Comorbidities** |  |  |
| Diabetes as cause of ESKD | 78,524 (38.2%) | 17,703 |
| Carpal tunnel syndrome | 8,091 (4.4%) | 37,630 |
| Dementia | 8,915 (5.0%) | 45,541 |
| Ischemic heart disease | 16,055 (8.6%) | 37,034 |
| Cerebral hemorrhage | 10,099 (5.4%) | 37,336 |
| Cerebral infarction | 30,588 (16.4%) | 36,393 |
| Limb amputation | 6,193 (3.3%) | 35,776 |
| Hip fracture | 6,563 (3.5%) | 37,530 |
| **Outcome** |  |  |
| Non-frail | 60,800 (27.2%) |  |
| Frail | 18,817 (8.4%) |  |
| Bedridden | 5,332 (2.4%) |  |
| Deceased | 138,552 (62.0%) |  |

Abbreviations: BMI, body mass index; Hb, hemoglobin; Ca, calcium; P, phosphorus; PTH, parathyroid hormone; CRP, C-reactive protein; β2MG, beta-2 microglobulin; Kt/V, single-pool dialyzer clearance of urea normalized to distribution volume; nPCR, normalized protein catabolic rate; ESKD, end-stage kidney disease; RPGN, rapidly progressive glomerulonephritis.

### Eight-Year Transitions in Physical Function Status from Baseline

Figure 1 presents a Sankey diagram illustrating 8-year transitions in physical function status, including frailty. Among the 188,996 patients with available data at baseline and year 8, 28.9% (n = 54,559) were non-frail, 8.8% (n = 16,625) were frail, 2.4% (n = 4,620) were bedridden, and 59.9% (n = 113,192) had died. Of the 137,947 patients who were non-frail at baseline, a substantial proportion (38.5%, n = 53,155) remained non-frail, while nearly half (49.1%, n = 67,708) had died. A notable proportion (10.1%, n = 13,864) progressed to frailty, and a small proportion (2.3%, n = 3,220) became bedridden. Among the 39,191 patients classified as frail at baseline, a modest proportion (6.7%, n = 2,626) remained frail, whereas the majority (86.9%, n = 34,050) had died. A small proportion either worsened to bedridden (3.0%, n = 1,159) or improved to non-frail status (3.5%, n = 1,356). For the 11,858 patients who were bedridden at baseline, a minimal proportion (2.0%, n = 241) remained bedridden, while nearly all (96.4%, n = 11,434) had died. Reversions to frail (1.1%, n = 135) or non-frail (0.4%, n = 48) were rare.

**Figure 1.**
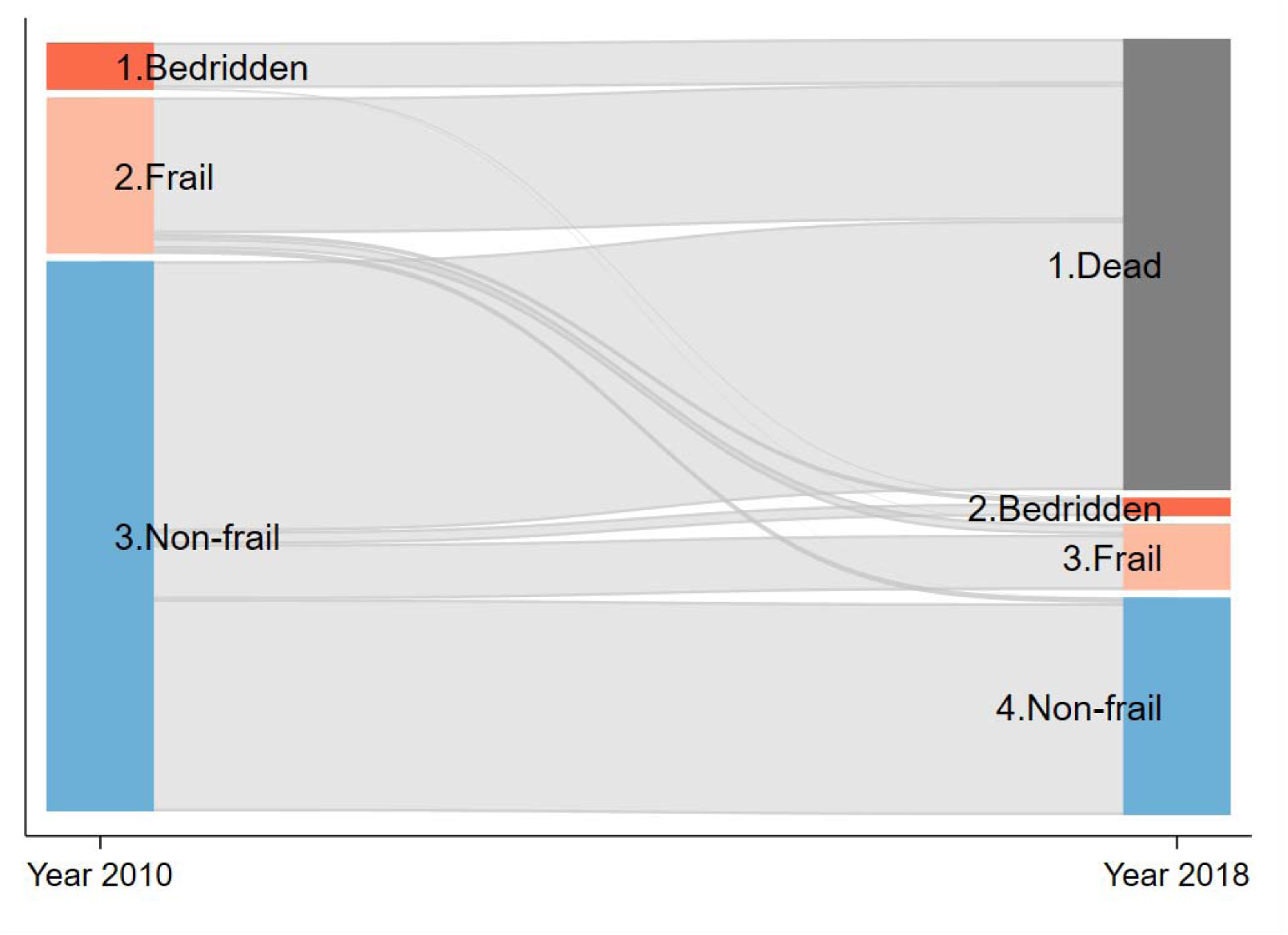
Eight-Year Transitions in Physical Function Status (n = 188,996) Sankey diagram showing transitions in physical function status between 2010 and 2018. It includes only individuals with complete data at both time points. Node height corresponds to the relative proportion of individuals in each status category at baseline (2010) and follow-up (2018). Flow width indicates the proportion of individuals transitioning between categories over time. Colors indicate status: blue = non-frail, light orange = frail, orange = bedridden, gray = deceased.

### Association of 8-Year Outcomes with Baseline Dialysis Duration and Physical Function (Relative Measures)

Table 2 presents the ORs of physical function outcomes at 8 years, stratified by baseline dialysis duration and physical function status. ORs for other predictors are shown in Supplementary Table 3. Dialysis duration demonstrated a dose-dependent association across all physical function outcomes.

**Table 2.** Associations between baseline dialysis duration, frailty status, and 8-year physical function outcomes.

| Variables | Non-frail |  | Frail |  | Bedridden |  | Deceased |  |
| --- | --- | --- | --- | --- | --- | --- | --- | --- |
|  | aOR (95%CI) | AME (95%CI) | aOR (95%CI) | AME (95%CI) | aOR (95%CI) | AME (95%CI) | aOR (95%CI) | AME (95%CI) |
| Dialysis duration, yr |  |  |  |  |  |  |  |  |
| 0-<5 | Base outcome for multinomial logit model | Reference | Reference | Reference | Reference | Reference | Reference | Reference |
| 5-<10 |  | -3.8% (-4.2%, -3.5%) | 1.13 (1.08, 1.18) | -0.9% (-1.2%, -0.6%) | 1.18 (1.09, 1.27) | -0.2% (-0.4%, -0.05%) | 1.47 (1.42, 1.52) | 4.9% (4.5%, 5.4%) |
| 15-<20 |  | -4.3% (-4.7%, -3.8%) | 1.18 (1.12, 1.25) | -0.7% (-1.0%, -0.3%) | 1.09 (1.00, 1.20) | -0.4% (-0.6%, -0.3%) | 1.53 (1.47, 1.59) | 5.4% (4.9%, 5.9%) |
| 20-<30 |  | -7.6% (-8.3%, -7.0%) | 1.67 (1.55, 1.81) | 0.5% (-0.1%, 1.1%) | 1.22 (1.05, 1.42) | -0.7% (-0.9%, -0.4%) | 2.04 (1.93, 2.17) | 7.8% (7.0%, 8.6%) |
| 30≤ |  | -14.5% (-15.5%, -13.6%) | 3.18 (2.77, 3.65) | 1.6% (0.6%, 2.6%) | 2.51 (1.98, 3.17) | -0.3% (-0.8%, 0.1%) | 4.16 (3.71, 4.65) | 13.2% (12.0%, 14.5%) |
| Baseline frailty |  |  |  |  |  |  |  |  |
| Non-frail |  | Reference | Reference | Reference | Reference | Reference | Reference | Reference |
| Frail |  | -17.0% (-17.5%, -16.4%) | 3.43 (3.18, 3.69) | 0.04% (-0.4%, 0.4%) | 5.39 (4.90, 5.93) | 1.1% (0.9%, 1.4%) | 5.43 (5.10, 5.79) | 15.8% (15.2%, 16.4%) |
| Bedridden |  | -22.6% (-24.0%, -21.2%) | 3.38 (2.40, 4.76) | -5.1% (-5.8%, -4.5%) | 15.1 (10.8, 21.2) | 1.7% (1.1%, 2.3%) | 16.4 (12.1, 22.1) | 26.0% (24.6%, 27.4%) |

For frailty status, the ORs (95% CI) increased with longer dialysis duration: 1.13 (1.08–1.18) for 5 to <10 years, 1.18 (1.12–1.25) for 10 to <20 years, 1.67 (1.55–1.81) for 20 to <30 years, and 3.18 (2.77–3.65) for ≥30 years. For bedridden status, the ORs were 1.18 (1.09–1.27), 1.09 (1.00–1.20), 1.22 (1.05–1.42), and 2.51 (1.98–3.17) for the corresponding dialysis duration categories. Mortality risk also rose progressively, with ORs of 1.47 (1.42–1.52), 1.53 (1.47–1.59), 2.04 (1.93–2.17), and 4.16 (3.71–4.65).

Baseline physical function status showed dose-dependent associations with outcomes at 8 years. For frailty status, the ORs were 3.43 (3.18–3.69) for baseline frailty and 3.38 (2.40– 4.76) for baseline bedridden. For bedridden status, the ORs were 5.39 (4.90–5.93) for baseline frailty and 15.1 (10.8–21.2) for baseline bedridden. For mortality status, the ORs were 5.43 (5.10–5.79) and 16.4 (12.1–22.1) for baseline frailty and baseline bedridden, respectively.

Diabetes, hypoalbuminemia, elevated CRP, a low creatinine index, and residence in institutional settings, such as nursing homes or hospitals, were all associated with frailty, bedridden status, and death at 8 years.

### Predicted Eight-Year Outcomes and Absolute Measure Associations by Baseline Dialysis Duration and Physical Function

Figure 2A shows the predicted probabilities of physical function outcomes at eight years, stratified by baseline dialysis duration. As dialysis duration increased from <5 years to ≥30 years, the predicted probability of death progressively rose from 58.8% to 72.0%, whereas the probability of being non-frail declined from 30.0% to 15.5%. The prevalence of frailty was lowest among individuals with 5 to <10 years of dialysis (7.8%) and gradually increased to 10.3% among those with ≥30 years of dialysis. Predicted probabilities for being bedridden varied minimally, with a slight decrease from 2.6% for <5 years to 1.9% for 20 to <30 years. Table 2 summarizes the differences by dialysis duration using AMEs, with <5 years serving as the reference. For non-frail status, AMEs (95% CI) steadily declined from −3.8% (−4.2%, −3.5%) for 5 to <10 years to −14.5% (−15.5%, −13.6%) for ≥30 years. In the case of frailty, AMEs ranged from −0.9% (−1.2%, −0.6%) for 5 to <10 years and −0.7% (−1.0%, −0.3%) for 10 to <20 years, then increased to 0.5% (−0.1%, 1.1%) for 20 to <30 years and 1.6% (0.6%, 2.6%) for ≥30 years. Bedridden status showed AMEs of −0.2% (−0.4%, −0.05%), −0.4% (−0.6%, −0.3%), −0.7% (−0.9%, −0.4%), and −0.3% (−0.8%, 0.1%), without a directional trend. The AMEs for mortality (95% CI) progressively increased from 4.9% (4.5%, 5.4%), 5.4% (4.9%, 5.9%), 7.8% (7.0%, 8.6%), to 13.2% (12.0%, 14.5%) for each respective dialysis duration category.

**Figure 2.**
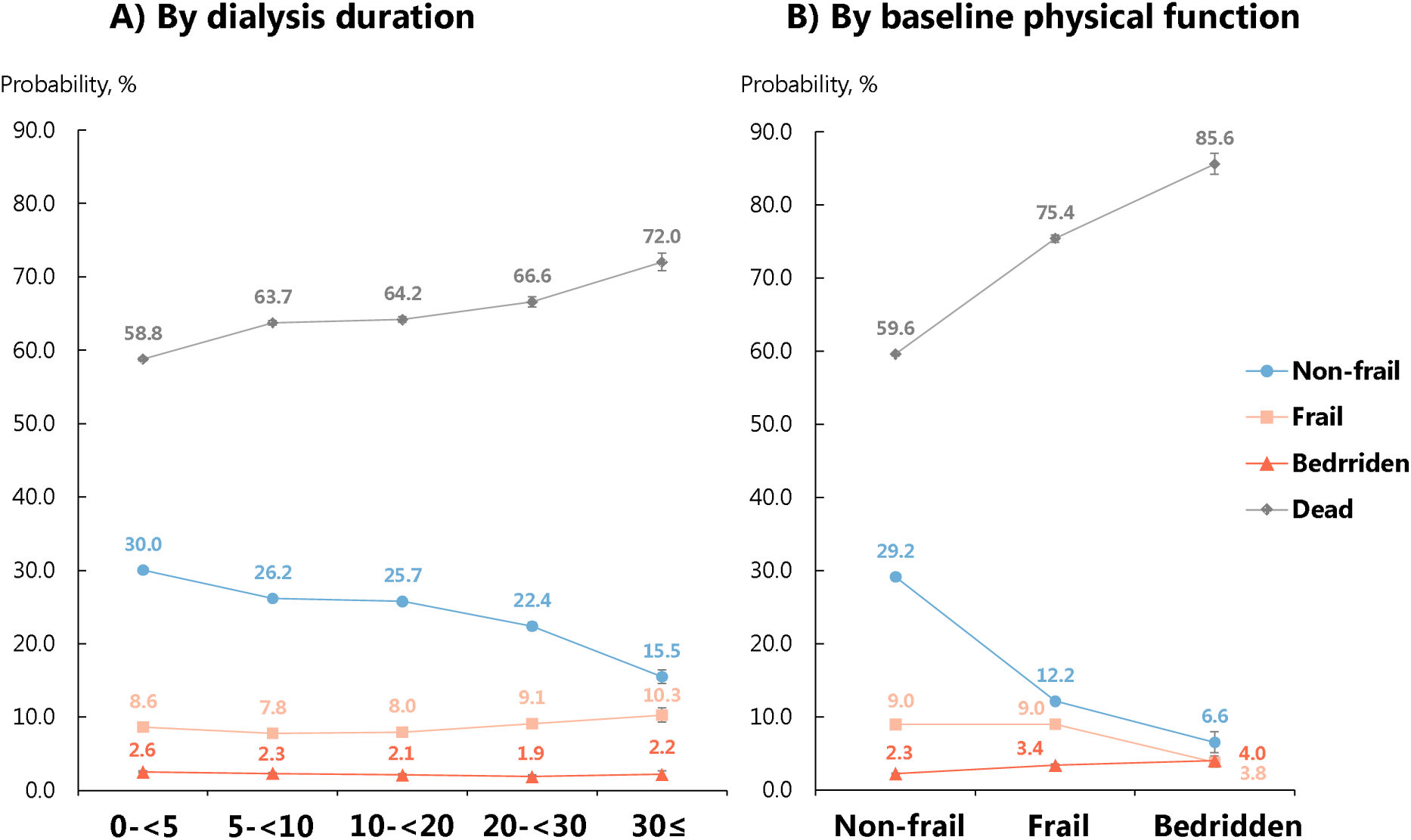
Predicted 8-Year Outcomes by Baseline Dialysis Duration and 545 Physical Function Status Predicted probabilities of 8-year physical function outcomes by (A) baseline dialysis duration and (B) physical function status. Outcomes were categorized as non-frail, frail, bedridden, or deceased, based on a multinomial logistic regression model. Baseline dialysis duration, physical function status, and all covariates listed in Table 1 were included in the model.

Figure 2B shows predicted probabilities of physical function outcomes at eight years, stratified by baseline physical function status. As baseline status progressed from non-frail to bedridden, the predicted probability of death substantially increased from 59.6% to 85.6%. By contrast, the probability of remaining non-frail markedly decreased from 29.2% to 6.6%.

Frailty probabilities peaked at baseline for non-frail and frail (9.0%) and substantially decreased for bedridden (3.8%). Predicted probabilities of being bedridden slightly increased from 2.3% for non-frail to 4.0% for bedridden.

The differences by baseline physical function outcomes are summarized in Table 2 using AMEs, with non-frail serving as the reference. For non-frail status, AMEs (95% CI) progressively decreased from −17.0% (−17.5%, −16.4%) for frail to −22.6% (−24.0%, −21.2%) for bedridden. For frailty status, AMEs were 0.04% (−0.4%, 0.4%) for frail and −5.1% (−5.8%, −4.5%) for bedridden. For bedridden status, AMEs slightly increased from 1.1% (0.9%, 1.4%) for frail to 1.7% (1.1%, 2.3%) for bedridden. The AME for death (95% CI) increased from 15.8% (15.2%, 16.4%) for frail to 26.0% (24.6%, 27.4%) for bedridden.

## Discussion

To the best of our knowledge, this study is the first to delineate long-term transitions in physical function status, including frailty, and examined their associations with baseline physical function and dialysis duration, using a nationwide cohort of patients undergoing maintenance HD. Due to the death of a slight majority (approximately 60%) of patients, only a modest proportion (<30%) survived in a non-frail state. The remaining patients included a minority who were frail and a minimal proportion who were bedridden. Longer dialysis duration was associated with a dose-dependent increase in the relative risk of frailty and bedridden status. However, absolute differences in physical function outcomes by dialysis duration were minimal, whereas mortality showed a notable variation. Similarly, poorer baseline physical function was associated with a dose-dependent increase in the relative risk of frailty, being bedridden, and death. In absolute terms, lower baseline physical function significantly increased mortality, whereas differences in frailty across baseline physical function levels remained minor.

The obtained findings on transitions in physical function, including frailty and bedridden status, both contrast with and add to the existing literature. First, unlike a two-year U.S. study in which similar proportions of patients improved or worsened in Fried frailty phenotype scores,^9^ very few patients in our cohort improved from frail or bedridden states. This discrepancy may be partially attributed to differences in follow-up duration and patient characteristics. Our study followed patients for 8 years, well beyond the average dialysis survival of less than 5 years.^26^ Moreover, our cohort had a mean age approximately 10 years older and a dialysis vintage about two years longer. In comparison, a study from Hong Kong with a median follow-up of nearly 4 years found that about one-fourth of PD patients transitioned from mildly frail to non-frail status^10^; however, their cohort was also roughly 10 years younger on average. Second, to our knowledge, this study is the first to demonstrate a dose-dependent association between dialysis duration and the incidence of frailty or bedridden status. Previous research, including our own, largely relied on cross-sectional designs to explore these relationships.^2,27^ By contrast, our findings on the association between longer dialysis duration and increased mortality are consistent with those of several previous studies.^28,29^ Third, this study provides novel evidence of dose-dependent risks for progression from baseline frailty to more severe outcomes, such as becoming bedridden or dying, an area previously underexplored. For instance, a previous study in PD patients found no association between baseline frailty scores and subsequent worsening frailty.^10^ While the prognostic value of frailty for mortality in end-stage kidney disease is well established,^30^ progression to bedridden status has not been reported. Fourth, previous studies with limited follow-up durations have not fully captured the disproportionately high number of deaths relative to transitions to frailty or bedridden status among patients with impaired physical function at baseline. For example, in a study of nursing home residents initiating dialysis, only 13% maintained functional status within one year, whereas 58% died and 29% experienced functional decline.^31^ In our 8-year follow-up, only 9.7% of patients classified as frail at baseline maintained or improved their status, while the vast majority (86.9%) died.

We believe these findings hold important clinical implications for dialysis care providers and researchers. First, the observed association between dialysis vintage and the progression to frailty or a bedridden state highlights the need to investigate unmeasured factors associated with long-term dialysis. These may include complications, such as dialysis-associated amyloidosis (e.g., destructive spondyloarthropathy), mineral and bone disorders, and protein-energy wasting, as well as their underlying mechanisms.^2^ Concurrently, this study confirmed previously reported factors associated with transitions from non-frail to frail or bedridden status, including diabetes,^9^ hypoalbuminemia,^9^ systemic inflammation (elevated CRP),^9,10^ malnutrition (low creatinine index),^10^ and hospitalizations.^9,10^

Second, our findings on long-term transitions in physical function can aid clinicians in strengthening shared decision-making regarding dialysis initiation and withdrawal. For example, even among patients who were non-frail at baseline, only approximately 40% remained non-frail after 8 years. These findings highlight the importance of preparing for future functional decline or death based on a patient’s current physical function status.

Patients and caregivers, regardless of frailty status, should be encouraged to engage in advance care planning aligned with personal values and goals, fostering a dialysis experience with fewer regrets. In addition to efforts to prevent transitions to frailty or bedridden states, greater attention may be warranted for preventing acute life-threatening events, such as cardiovascular disease and infections, which may exert a larger absolute impact on mortality. Finally, baseline frailty or bedridden status was increasingly associated with subsequent functional decline, the absolute risk of transitioning to death was substantially greater. Among patients who were frail at baseline, few remained in that state, while the majority died during the follow-up period. These findings emphasize the limited capacity of long-term dialysis to preserve physical function and underscore the poor prognosis often encountered by dialysis providers, patients, and their families. By providing a realistic outlook, these results may guide patients who consider dialysis despite a low probability of survival,^32^ and support discussions about dialysis initiation or palliative care among older, frail individuals who prioritize functional independence.^3^

This study has several strengths. First, the use of a nationwide registry including nearly all dialysis facilities in Japan provided sufficient statistical power to detect associations involving dialysis duration of ≥30 years, frailty, and bedridden status. Second, the study contributes valuable data on 8-year transitions in physical function within a dialysis cohort, where average survival is typically <5 years.^26^ Third, key confounders and intermediates related to physical function, such as age, nutritional status, dementia, β2MG, carpal tunnel syndrome, hip fracture, and residential setting, were accounted for, enhancing the validity of our adjusted findings in elucidating the mechanisms linking long-term dialysis and baseline physical function to future frailty and bedridden status.

Several limitations should be acknowledged. First, frailty was assessed using the clinician-rated ECOG PS scale rather than more objective measures such as the Fried criteria, which includes measures like gait speed and grip strength.^33^ However, collecting such data across a nationwide cohort is impractical, as these assessments are not routinely performed in daily clinical practice. As in our previous and other large-scale studies, the ECOG PS-based definition was adopted to ensure the feasibility of the study.^2,17^ Second, although the study captured long-term transitions in frailty status, more detailed trajectories could not be evaluated. For example, among those transitioning from non-frail to death, identifying those who may have experienced intermediate stages of frailty or bedridden status was not possible.

Similarly, among those classified as frail at both baseline and follow-up, transient recovery to a non-frail state could not be assessed. These insights would require more granular assessments of physical function, ideally on an annual or more frequent basis. Third, unmeasured factors may have influenced our findings. For instance, destructive spondyloarthropathy, a potential complication of long-term dialysis, may serve as an intermediate factor linking dialysis duration to physical function decline and may also confound the relationship between baseline physical function and survival. Fourth, the study population consisted almost entirely of Japanese patients, limiting the generalizability of findings related to 8-year predicted mortality rates and AMEs to other countries. An international cohort study of HD patients previously reported a 2.8- to 3.8-fold difference in mortality rates among Japan, the United States, and Europe.^34^ However, given that more than two decades have passed since that report, further investigation is needed to assess the applicability of our findings to dialysis cohorts in other healthcare systems and among more racially diverse populations.

In this nationwide 8-year follow-up study of Japanese HD patients, a significant proportion (28.9%) remained non-frail, while a modest proportion (8.8%) progressed to frailty and a minimal proportion (2.4%) became bedridden; more than half (59.9%) died during the follow-up period. Although long dialysis duration and baseline frailty or bedridden status were associated with subsequent deterioration in physical function, the absolute differences observed were relatively small. Further studies using more granular data on physical function are needed to provide patients with clearer insights into the specific trajectories of physical function decline.

## Disclosure

NK has received consulting fees from GlaxoSmithKline K.K. and Kyowa Kirin Co., Ltd. He has also received honoraria for speaking engagements and educational events from Eisai Co., Ltd., Taisho Pharmaceutical Co., Ltd., Kyowa Kirin Co., Ltd., GlaxoSmithKline K.K., Takeda Pharmaceutical Co., Ltd., Kissei Pharmaceutical Co., Ltd., and Vantive Japan. RI has received honoraria for speaking engagements and educational events from Astellas Pharma Inc., Novartis Pharma K.K., Otsuka Pharmaceutical and Vantive Japan. TT has received consulting fees from Astellas Pharma Inc. He has also honoraria for speaking engagements and educational events from Torii Pharmaceutical Co., Ltd., Ono Pharmaceutical Co., Ltd., Kyowa Kirin Co., Ltd., AstraZeneca K.K., Kissei Pharmaceutical Co., Ltd., and Nobelpharma Co., Ltd. MA has received payment for speaking from Novartis Pharma K.K., Otsuka Pharmaceutical. SY has received honoraria and consulting fee from Sanwa Kagaku Kenkyusho Co., Ltd. (SKK); honoraria from Kyowa Kirin Co., Ltd., Kissei Pharmaceutical Co., Ltd., Ono Pharmaceutical Co., Ltd., and Torii Pharmaceutical Co., Ltd.; and research findings from Toray Medical Co., Ltd. and Kaneka Medix Co., Ltd.

## Acknowledgements

We wish to acknowledge the efforts of the members of the subcommittee for JRDR Regional Cooperation and staff members of the dialysis facilities who participated in the survey and responded to the questionnaires. This study utilized data from the JRDR. The interpretation and reporting of these data are solely the responsibility of the authors and do not reflect the official views or policies of the JSDT.

## Data availability statement

Data will be available immediately after publication with no end dates. Data will be shared upon reasonable request to the corresponding author with permission from the JRDR investigators. Restrictions apply to the availability of the data analysed in this study to preserve patient confidentiality.

## Author contributions

Research idea and study design: KN, NK, SY; data acquisition: SY, TM; data analysis and interpretation: KN, NK, RI, TT, SY; statistical analysis: KN, NK; drafting of the manuscript: KN, NK; supervision or mentorship: SY, MA, TM. Each author contributed important intellectual content during manuscript drafting or revision, agreed to be personally accountable for the individual’s own contributions, and ensured that questions pertaining to the accuracy or integrity of any portion of the work, even one in which the author was not directly involved, were appropriately investigated and resolved, including documentation in the literature, if appropriate.

---

**Supplementary Figure 1.**
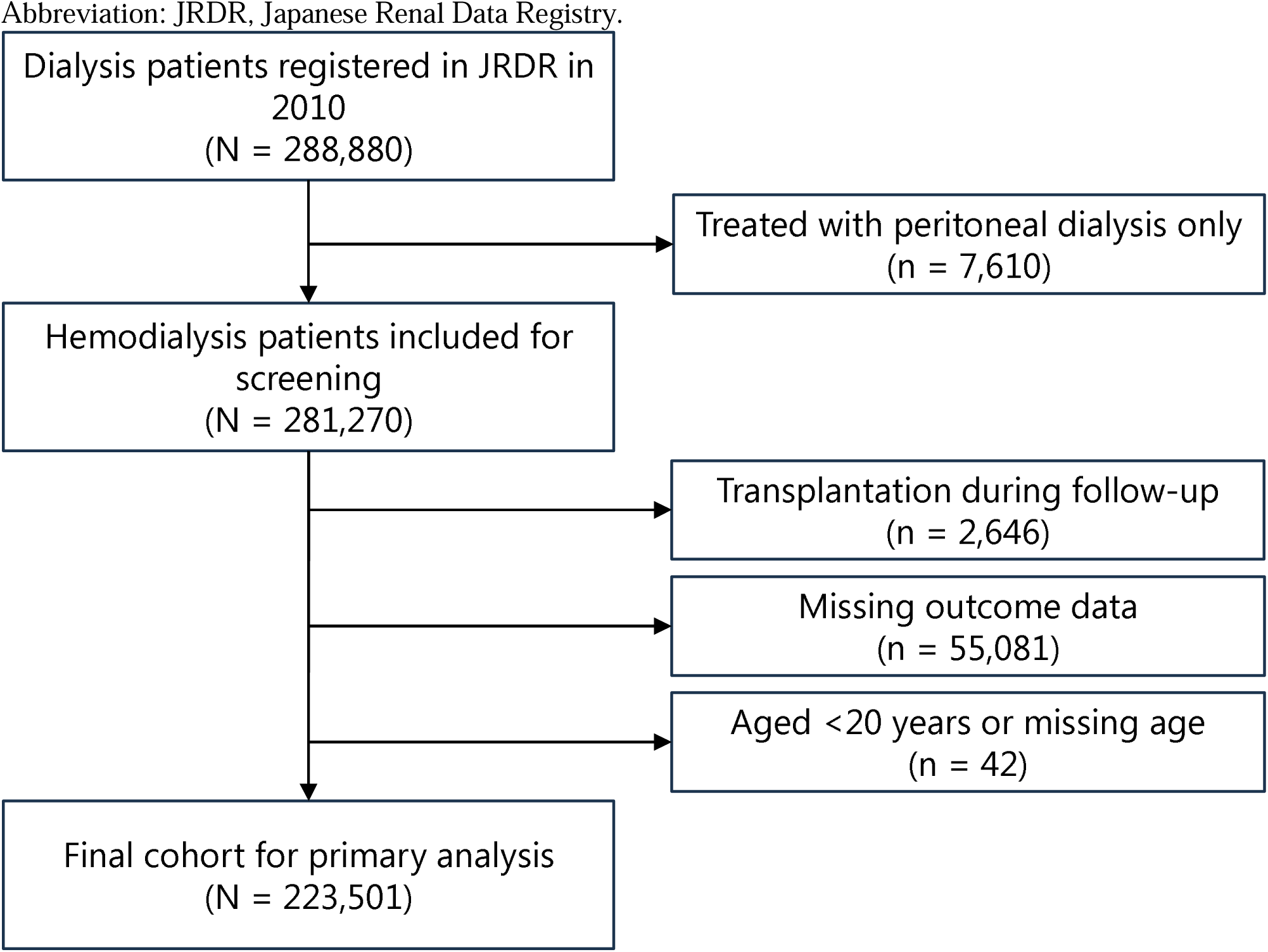
Participant Flow Diagram for the Primary Analysis Cohort

**Supplementary Table 1.**
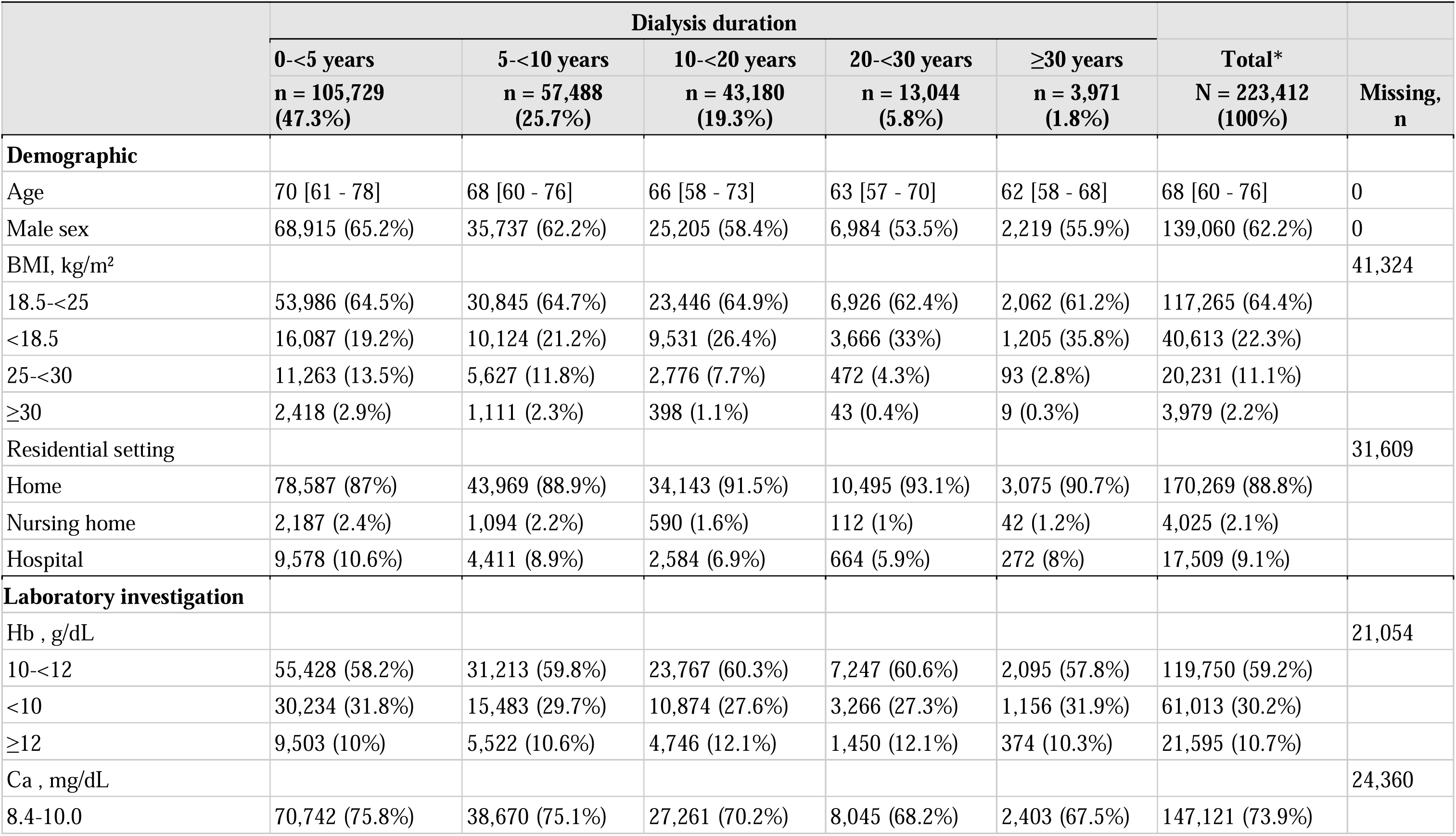
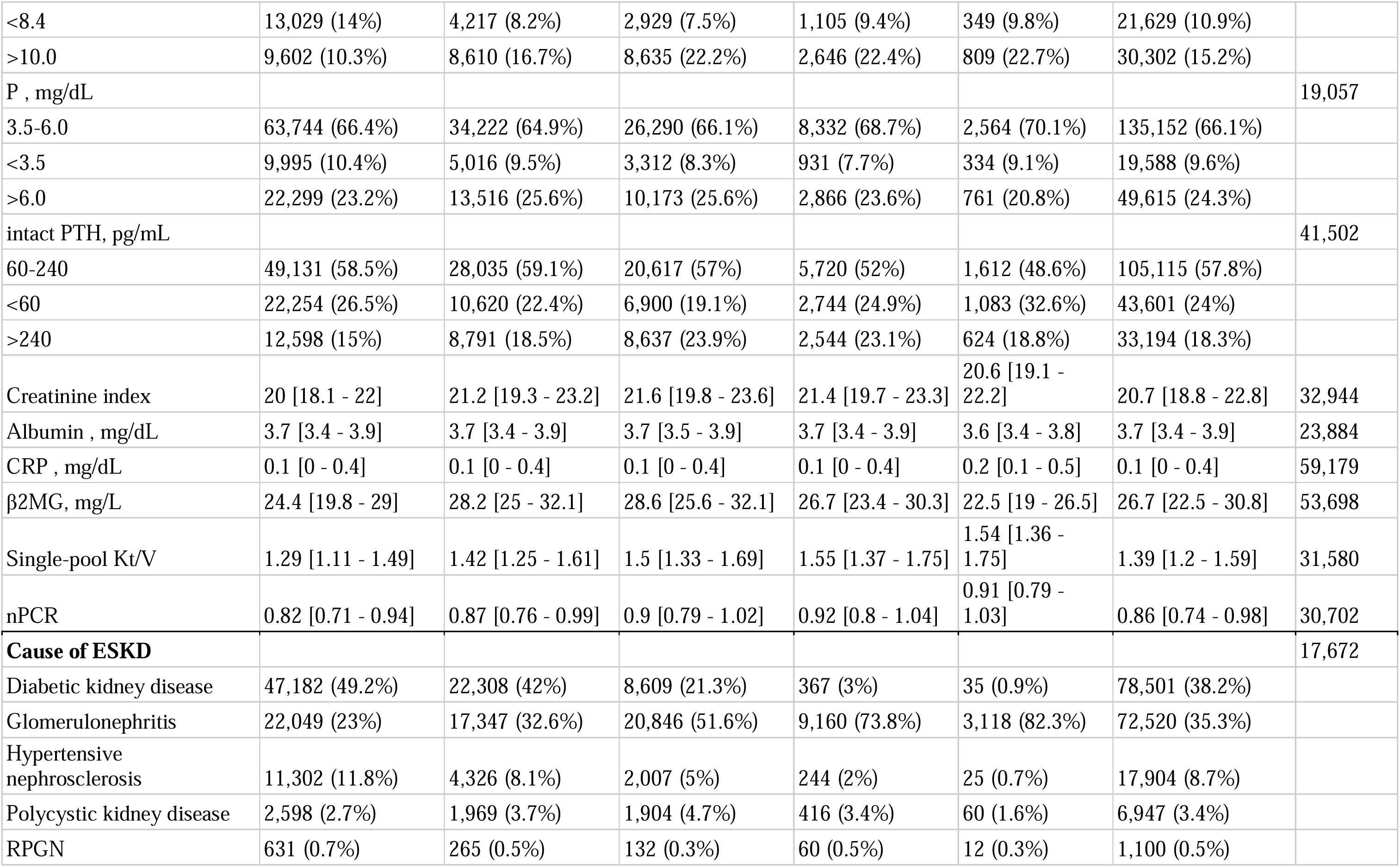
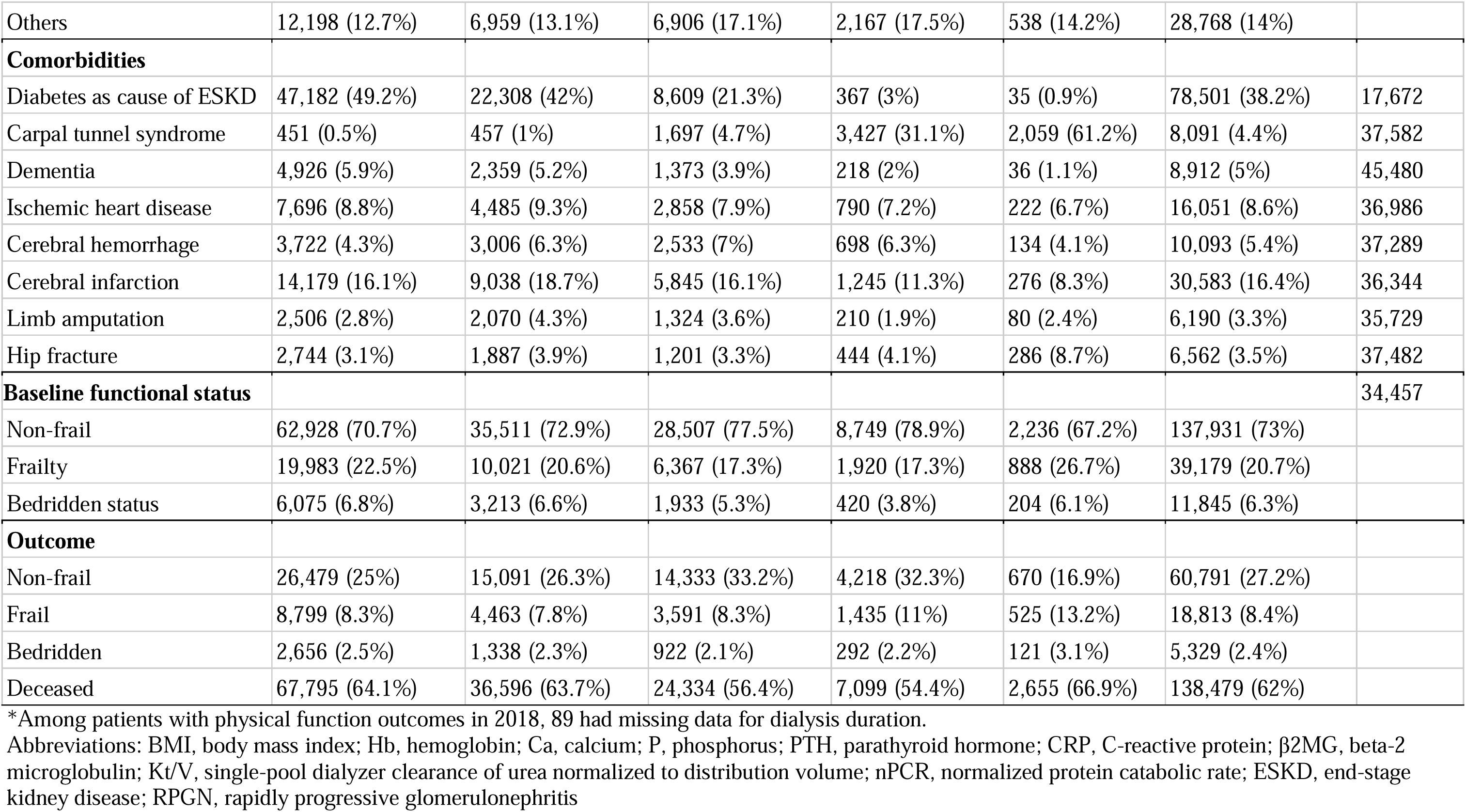
Baseline characteristics by dialysis duration (N = 223,412)Supplementary Table 2. Baseline characteristics by dialysis duration (N = 188,996)

|  | Dialysis duration |  |  |  |  | Total* | Missing,<br>n |
| --- | --- | --- | --- | --- | --- | --- | --- |
|  | 0-<5 years | 5-<10 years | 10-<20 years | 20-<30 years | ≥30 years |  |  |
|  | n = 105,729<br>(47.3%) | n = 57,488<br>(25.7%) | n = 43,180<br>(19.3%) | n = 13,044<br>(5.8%) | n = 3,971<br>(1.8%) | N = 223,412<br>(100%) |  |
| **Demographic** |  |  |  |  |  |  |  |
| Age | 70 [61 - 78] | 68 [60 - 76] | 66 [58 - 73] | 63 [57 - 70] | 62 [58 - 68] | 68 [60 - 76] | 0 |
| Male sex | 68,915 (65.2%) | 35,737 (62.2%) | 25,205 (58.4%) | 6,984 (53.5%) | 2,219 (55.9%) | 139,060 (62.2%) | 0 |
| BMI, kg/m<sup>2</sup> |  |  |  |  |  |  | 41,324 |
| 18.5-<25 | 53,986 (64.5%) | 30,845 (64.7%) | 23,446 (64.9%) | 6,926 (62.4%) | 2,062 (61.2%) | 117,265 (64.4%) |  |
| <18.5 | 16,087 (19.2%) | 10,124 (21.2%) | 9,531 (26.4%) | 3,666 (33%) | 1,205 (35.8%) | 40,613 (22.3%) |  |
| 25-<30 | 11,263 (13.5%) | 5,627 (11.8%) | 2,776 (7.7%) | 472 (4.3%) | 93 (2.8%) | 20,231 (11.1%) |  |
| ≥30 | 2,418 (2.9%) | 1,111 (2.3%) | 398 (1.1%) | 43 (0.4%) | 9 (0.3%) | 3,979 (2.2%) |  |
| Residential setting |  |  |  |  |  |  | 31,609 |
| Home | 78,587 (87%) | 43,969 (88.9%) | 34,143 (91.5%) | 10,495 (93.1%) | 3,075 (90.7%) | 170,269 (88.8%) |  |
| Nursing home | 2,187 (2.4%) | 1,094 (2.2%) | 590 (1.6%) | 112 (1%) | 42 (1.2%) | 4,025 (2.1%) |  |
| Hospital | 9,578 (10.6%) | 4,411 (8.9%) | 2,584 (6.9%) | 664 (5.9%) | 272 (8%) | 17,509 (9.1%) |  |
| **Laboratory investigation** |  |  |  |  |  |  |  |
| Hb , g/dL |  |  |  |  |  |  | 21,054 |
| 10-<12 | 55,428 (58.2%) | 31,213 (59.8%) | 23,767 (60.3%) | 7,247 (60.6%) | 2,095 (57.8%) | 119,750 (59.2%) |  |
| <10 | 30,234 (31.8%) | 15,483 (29.7%) | 10,874 (27.6%) | 3,266 (27.3%) | 1,156 (31.9%) | 61,013 (30.2%) |  |
| ≥12 | 9,503 (10%) | 5,522 (10.6%) | 4,746 (12.1%) | 1,450 (12.1%) | 374 (10.3%) | 21,595 (10.7%) |  |
| Ca , mg/dL |  |  |  |  |  |  | 24,360 |
| 8.4-10.0 | 70,742 (75.8%) | 38,670 (75.1%) | 27,261 (70.2%) | 8,045 (68.2%) | 2,403 (67.5%) | 147,121 (73.9%) |  |
| <8.4 | 13,029 (14%) | 4,217 (8.2%) | 2,929 (7.5%) | 1,105 (9.4%) | 349 (9.8%) | 21,629 (10.9%) |  |
| >10.0 | 9,602 (10.3%) | 8,610 (16.7%) | 8,635 (22.2%) | 2,646 (22.4%) | 809 (22.7%) | 30,302 (15.2%) |  |
| P , mg/dL |  |  |  |  |  |  | 19,057 |
| 3.5-6.0 | 63,744 (66.4%) | 34,222 (64.9%) | 26,290 (66.1%) | 8,332 (68.7%) | 2,564 (70.1%) | 135,152 (66.1%) |  |
| <3.5 | 9,995 (10.4%) | 5,016 (9.5%) | 3,312 (8.3%) | 931 (7.7%) | 334 (9.1%) | 19,588 (9.6%) |  |
| >6.0 | 22,299 (23.2%) | 13,516 (25.6%) | 10,173 (25.6%) | 2,866 (23.6%) | 761 (20.8%) | 49,615 (24.3%) |  |
| intact PTH, pg/mL |  |  |  |  |  |  | 41,502 |
| 60-240 | 49,131 (58.5%) | 28,035 (59.1%) | 20,617 (57%) | 5,720 (52%) | 1,612 (48.6%) | 105,115 (57.8%) |  |
| <60 | 22,254 (26.5%) | 10,620 (22.4%) | 6,900 (19.1%) | 2,744 (24.9%) | 1,083 (32.6%) | 43,601 (24%) |  |
| >240 | 12,598 (15%) | 8,791 (18.5%) | 8,637 (23.9%) | 2,544 (23.1%) | 624 (18.8%) | 33,194 (18.3%) |  |
| Creatinine index | 20 [18.1 - 22] | 21.2 [19.3 - 23.2] | 21.6 [19.8 - 23.6] | 21.4 [19.7 - 23.3] | 20.6 [19.1 - 22.2] | 20.7 [18.8 - 22.8] | 32,944 |
| Albumin , mg/dL | 3.7 [3.4 - 3.9] | 3.7 [3.4 - 3.9] | 3.7 [3.5 - 3.9] | 3.7 [3.4 - 3.9] | 3.6 [3.4 - 3.8] | 3.7 [3.4 - 3.9] | 23,884 |
| CRP , mg/dL | 0.1 [0 - 0.4] | 0.1 [0 - 0.4] | 0.1 [0 - 0.4] | 0.1 [0 - 0.4] | 0.2 [0.1 - 0.5] | 0.1 [0 - 0.4] | 59,179 |
| β2MG, mg/L | 24.4 [19.8 - 29] | 28.2 [25 - 32.1] | 28.6 [25.6 - 32.1] | 26.7 [23.4 - 30.3] | 22.5 [19 - 26.5] | 26.7 [22.5 - 30.8] | 53,698 |
| Single-pool Kt/V | 1.29 [1.11 - 1.49] | 1.42 [1.25 - 1.61] | 1.5 [1.33 - 1.69] | 1.55 [1.37 - 1.75] | 1.54 [1.36 - 1.75] | 1.39 [1.2 - 1.59] | 31,580 |
| nPCR | 0.82 [0.71 - 0.94] | 0.87 [0.76 - 0.99] | 0.9 [0.79 - 1.02] | 0.92 [0.8 - 1.04] | 0.91 [0.79 - 1.03] | 0.86 [0.74 - 0.98] | 30,702 |
| **Cause of ESKD** |  |  |  |  |  |  | 17,672 |
| Diabetic kidney disease | 47,182 (49.2%) | 22,308 (42%) | 8,609 (21.3%) | 367 (3%) | 35 (0.9%) | 78,501 (38.2%) |  |
| Glomerulonephritis | 22,049 (23%) | 17,347 (32.6%) | 20,846 (51.6%) | 9,160 (73.8%) | 3,118 (82.3%) | 72,520 (35.3%) |  |
| Hypertensive nephrosclerosis | 11,302 (11.8%) | 4,326 (8.1%) | 2,007 (5%) | 244 (2%) | 25 (0.7%) | 17,904 (8.7%) |  |
| Polycystic kidney disease | 2,598 (2.7%) | 1,969 (3.7%) | 1,904 (4.7%) | 416 (3.4%) | 60 (1.6%) | 6,947 (3.4%) |  |
| RPGN | 631 (0.7%) | 265 (0.5%) | 132 (0.3%) | 60 (0.5%) | 12 (0.3%) | 1,100 (0.5%) |  |
| Others | 12,198 (12.7%) | 6,959 (13.1%) | 6,906 (17.1%) | 2,167 (17.5%) | 538 (14.2%) | 28,768 (14%) |  |
| **Comorbidities** |  |  |  |  |  |  |  |
| Diabetes as cause of ESKD | 47,182 (49.2%) | 22,308 (42%) | 8,609 (21.3%) | 367 (3%) | 35 (0.9%) | 78,501 (38.2%) | 17,672 |
| Carpal tunnel syndrome | 451 (0.5%) | 457 (1%) | 1,697 (4.7%) | 3,427 (31.1%) | 2,059 (61.2%) | 8,091 (4.4%) | 37,582 |
| Dementia | 4,926 (5.9%) | 2,359 (5.2%) | 1,373 (3.9%) | 218 (2%) | 36 (1.1%) | 8,912 (5%) | 45,480 |
| Ischemic heart disease | 7,696 (8.8%) | 4,485 (9.3%) | 2,858 (7.9%) | 790 (7.2%) | 222 (6.7%) | 16,051 (8.6%) | 36,986 |
| Cerebral hemorrhage | 3,722 (4.3%) | 3,006 (6.3%) | 2,533 (7%) | 698 (6.3%) | 134 (4.1%) | 10,093 (5.4%) | 37,289 |
| Cerebral infarction | 14,179 (16.1%) | 9,038 (18.7%) | 5,845 (16.1%) | 1,245 (11.3%) | 276 (8.3%) | 30,583 (16.4%) | 36,344 |
| Limb amputation | 2,506 (2.8%) | 2,070 (4.3%) | 1,324 (3.6%) | 210 (1.9%) | 80 (2.4%) | 6,190 (3.3%) | 35,729 |
| Hip fracture | 2,744 (3.1%) | 1,887 (3.9%) | 1,201 (3.3%) | 444 (4.1%) | 286 (8.7%) | 6,562 (3.5%) | 37,482 |
| **Baseline functional status** |  |  |  |  |  |  | 34,457 |
| Non-frail | 62,928 (70.7%) | 35,511 (72.9%) | 28,507 (77.5%) | 8,749 (78.9%) | 2,236 (67.2%) | 137,931 (73%) |  |
| Frailty | 19,983 (22.5%) | 10,021 (20.6%) | 6,367 (17.3%) | 1,920 (17.3%) | 888 (26.7%) | 39,179 (20.7%) |  |
| Bedridden status | 6,075 (6.8%) | 3,213 (6.6%) | 1,933 (5.3%) | 420 (3.8%) | 204 (6.1%) | 11,845 (6.3%) |  |
| **Outcome** |  |  |  |  |  |  |  |
| Non-frail | 26,479 (25%) | 15,091 (26.3%) | 14,333 (33.2%) | 4,218 (32.3%) | 670 (16.9%) | 60,791 (27.2%) |  |
| Frail | 8,799 (8.3%) | 4,463 (7.8%) | 3,591 (8.3%) | 1,435 (11%) | 525 (13.2%) | 18,813 (8.4%) |  |
| Bedridden | 2,656 (2.5%) | 1,338 (2.3%) | 922 (2.1%) | 292 (2.2%) | 121 (3.1%) | 5,329 (2.4%) |  |
| Deceased | 67,795 (64.1%) | 36,596 (63.7%) | 24,334 (56.4%) | 7,099 (54.4%) | 2,655 (66.9%) | 138,479 (62%) |  |

\*Among patients with physical function outcomes in 2018, 89 had missing data for dialysis duration.
Abbreviations: BMI, body mass index; Hb, hemoglobin; Ca, calcium; P, phosphorus; PTH, parathyroid hormone; CRP, C-reactive protein; β2MG, beta-2 microglobulin; Kt/V, single-pool dialyzer clearance of urea normalized to distribution volume; nPCR, normalized protein catabolic rate; ESKD, end-stage kidney disease; RPGN, rapidly progressive glomerulonephritis

**Supplementary Table 2.**
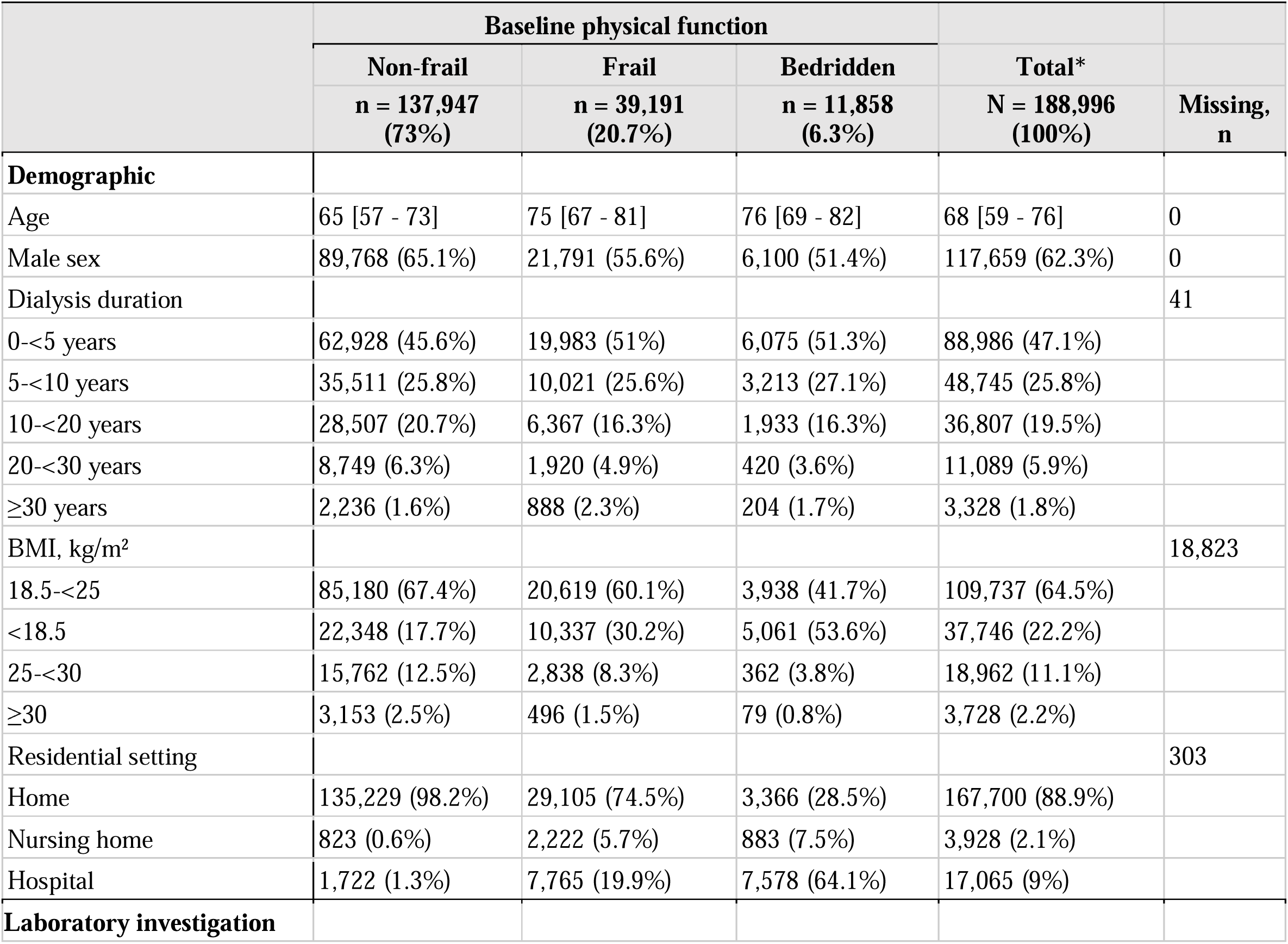
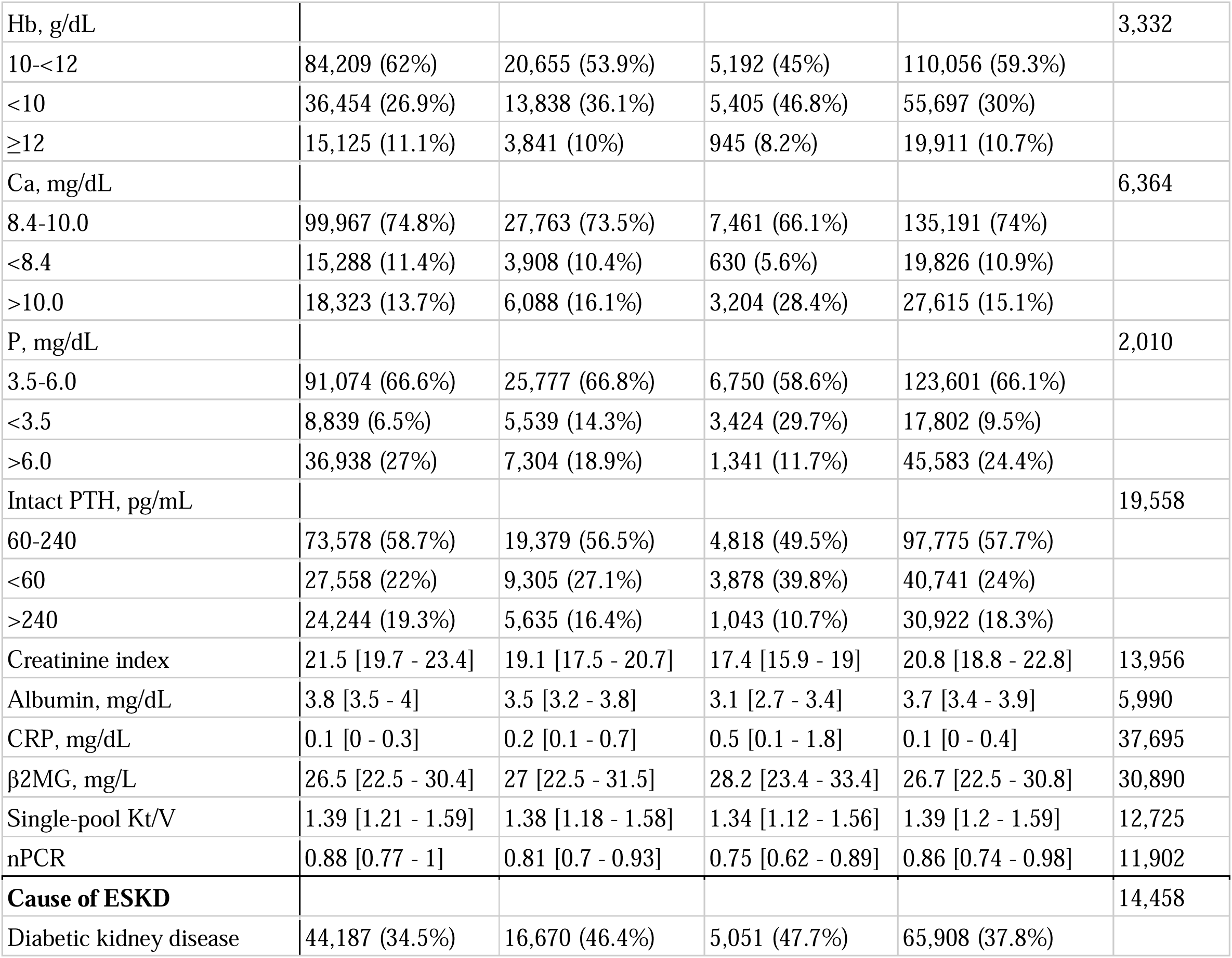
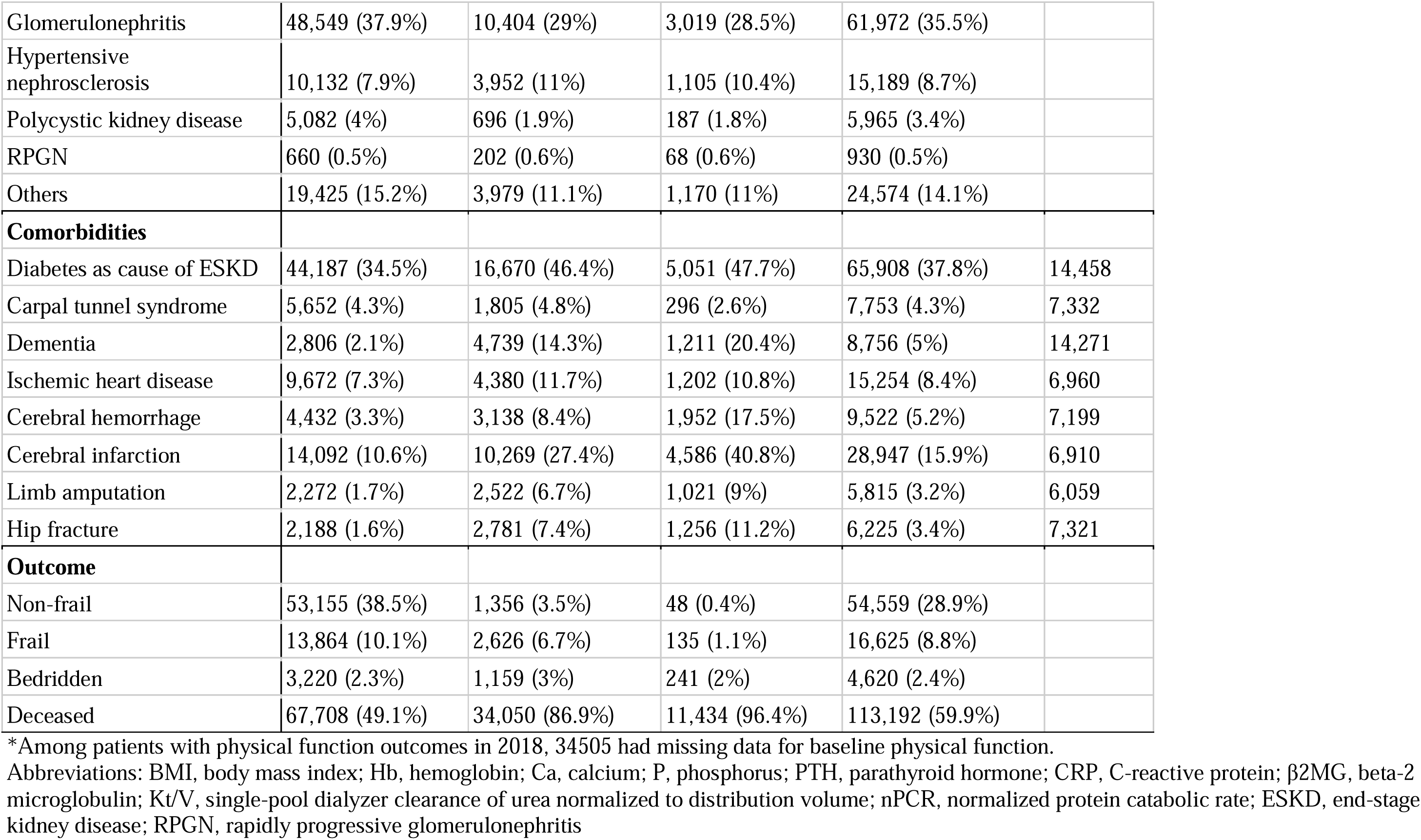
Baseline characteristics by dialysis duration (N = 188,996)

**Supplementary Table 3.**
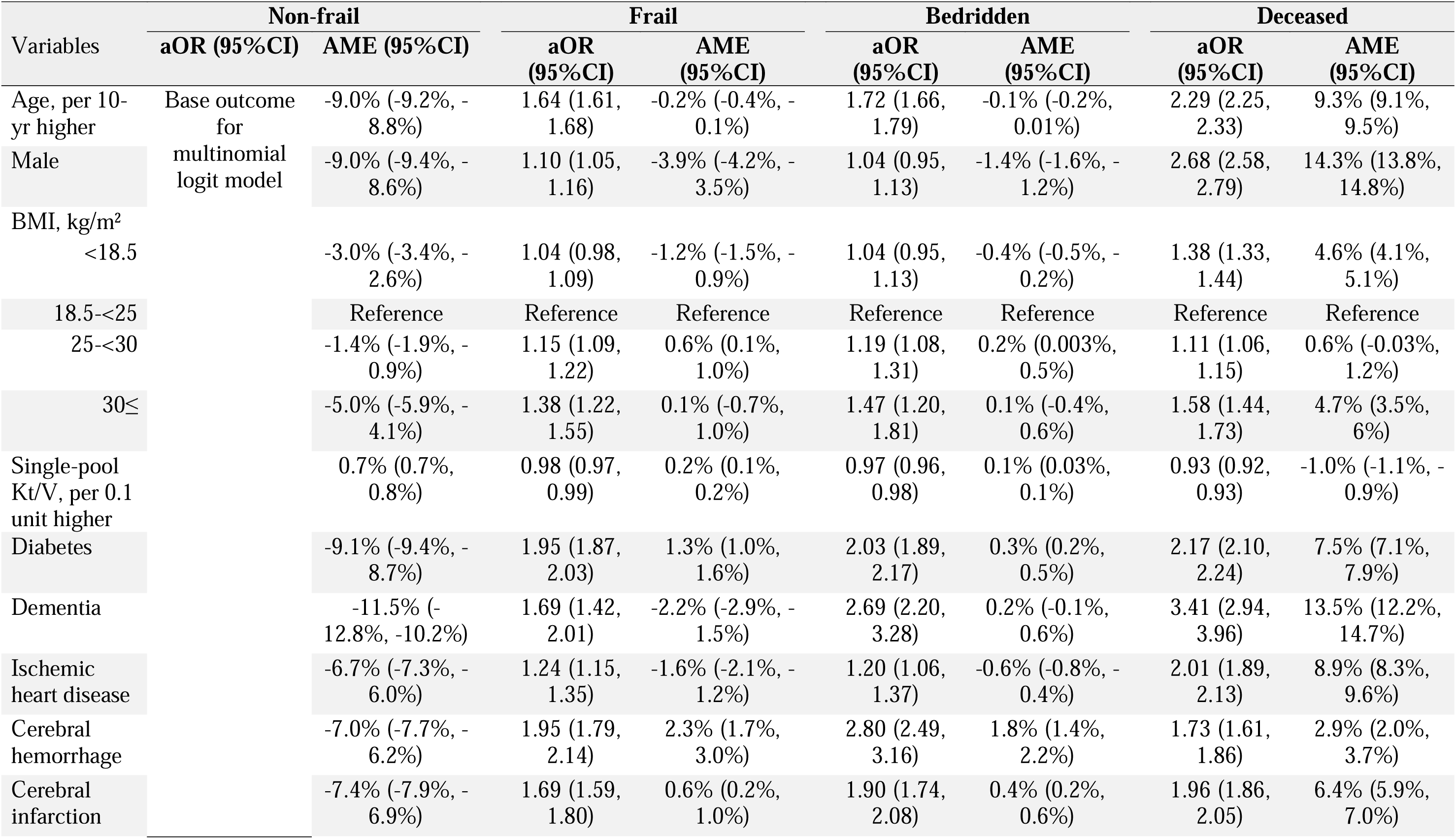
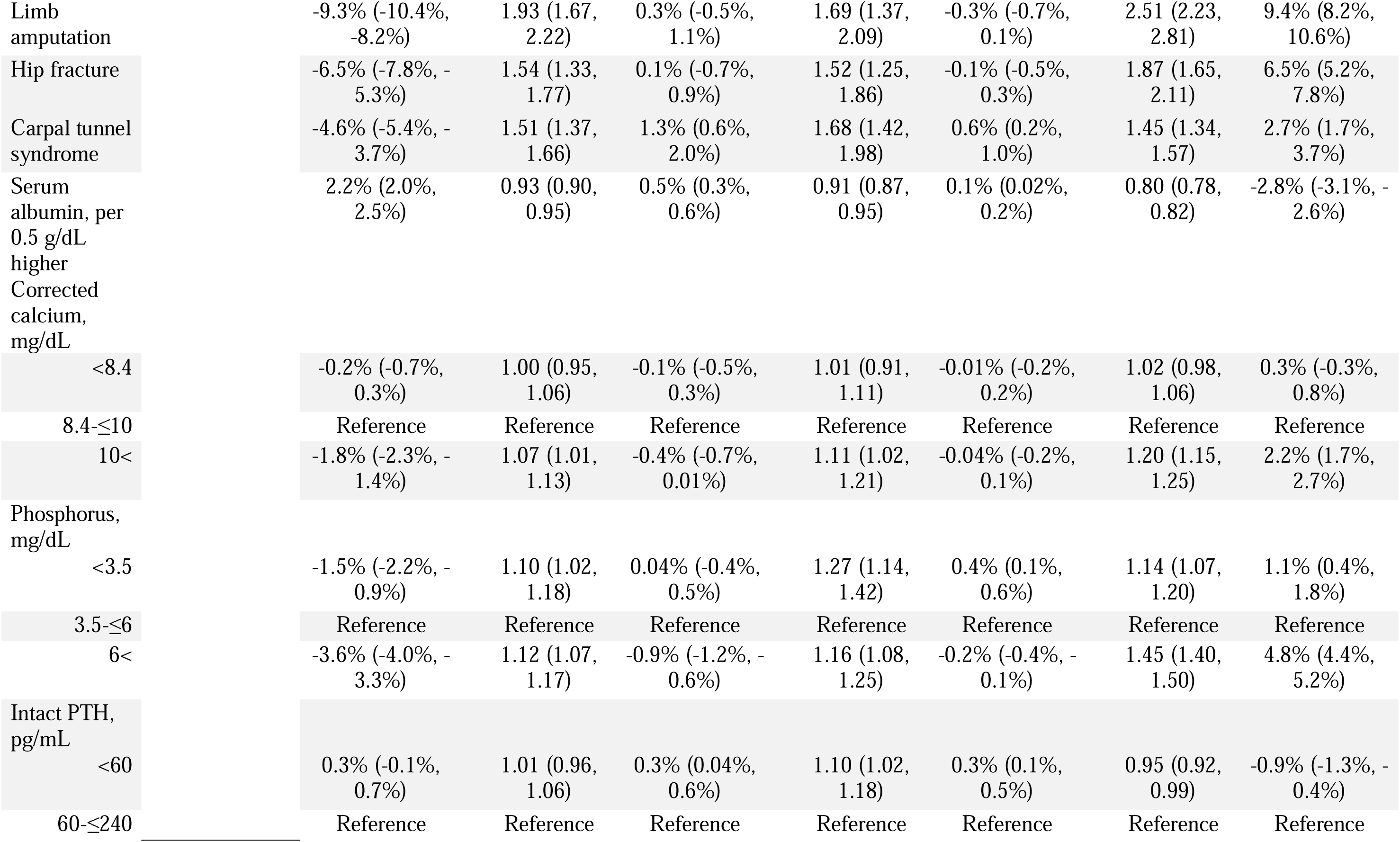
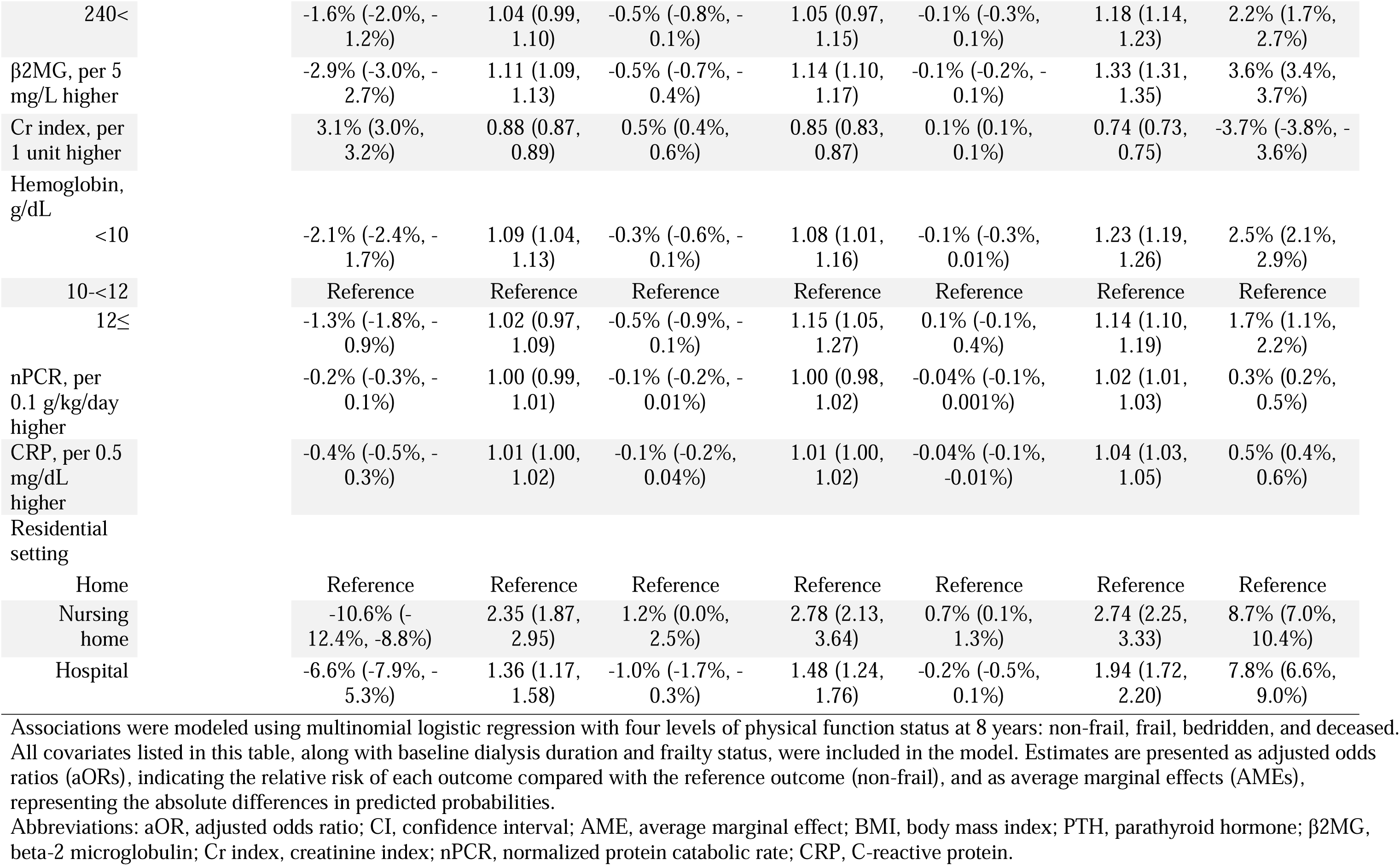
Associations between baseline covariates and 8-year physical function status.

